# Asthma severity and corticosteroid response depend on variable type 1 and type 2 inflammation in the airway

**DOI:** 10.1101/2023.10.05.23296609

**Authors:** John V. Fahy, Nathan D. Jackson, Satria P. Sajuthi, Elmar Pruesse, Camille M. Moore, Jamie L. Everman, Cydney Rios, Monica Tang, Marc Gauthier, Sally E. Wenzel, Eugene R. Bleecker, Mario Castro, Suzy A. Comhair, Serpil C. Erzurum, Annette T. Hastie, Wendy Moore, Elliot Israel, Bruce D. Levy, Loren Denlinger, Nizar N. Jarjour, Mats W. Johansson, David T. Mauger, Brenda R. Phillips, Kaharu Sumino, Prescott G. Woodruff, Michael C. Peters, Max A. Seibold, the National Heart, Lung, and Blood Institute Severe Asthma Research Program-3

## Abstract

The prevalence, inter-relationships, and longitudinal behavior of type 1 (T1) and type 2 (T2) immune responses in asthma are uncertain, as is the role of viruses as determinants of these responses. Here, we performed whole transcriptome network analysis on sputum cells collected from Severe Asthma Research Program (SARP)-3 patients before and after treatment with intramuscular corticosteroid and again at 1 and 3-year follow-up visits. We used network analysis to analyze whole-transcriptome gene expression and metagenomic analysis of these RNA-seq data to detect viruses. We identified T1 and T2 airway networks, the expression of which showed that 26% and 44% of patients had T1-high and T2-high asthma at baseline, respectively. Asthma severity outcomes were worse in T2-high asthma than in T1-high asthma and most severe in the subgroup of patients (14%) with combined T1- and T2-high disease. Corticosteroid treatment suppressed T2 but not T1 gene expression, and corticosteroid-associated improvements in FEV1 occurred only in patients with T1-L/T2-H disease and not in T1-H/T2-H patients. Although T1 and T2 inflammation at baseline was a significant predictor of T1 and T2 inflammation at follow-up visits, most patients had variable rather than persistent expression of T1 and T2 network genes. Viral metagenomic analyses uncovered that 24% of asthma sputum samples tested positive for a virus and high viral carriage was associated with an 11-fold increased risk of T1-high disease. Together our results uncover a relatively high burden of T1-high and T1/T2-high disease subtypes in severe asthma, which are corticosteroid-resistant and manifest with sub-clinical viral infection.

## INTRODUCTION

The clinical heterogeneity of asthma is explained by variability in underlying cellular and molecular mechanisms and the classification of asthma using its specific molecular features (endotyping) has evolved to inform precision medicine treatments. The best established asthma endotype is type 2-high asthma which describes patients who have upregulated airway expression of type 2 (T2) cytokines (IL-4, IL-5, and IL- 13) and increased airway infiltration by eosinophils and mast cells (*1, 2*). Milder forms of T2-high asthma, often occurring in younger and atopic patients, are usually responsive to corticosteroid treatment (*3*), but more severe T2-high disease, usually in older patients who frequently report sinusitis and nasal polyps as co-morbidities, are less corticosteroid responsive (*4–6*). The reasons for differing steroid responses in these patients are not known. Similarly, recently developed therapeutic proteins, which block ligands and receptors in the T2 inflammatory cascade, are clinically effective in many but not all patients with T2-high disease (*7–10*). Further complicating our understanding and treatment of the T2 endotype is a lack of data regarding the longitudinal persistence of T2 inflammation within patients. The identification of multiple T2 pathway genes harboring asthma risk genetic variants suggests that the propensity to airway T2 inflammation is an inherited patient-level characteristic (*11, 12*) that is molecularly/cellularly “wired” into the airway of a patient. Yet the invasiveness of airway sampling has prevented the repeated, longitudinal collection of patient airway specimens required to investigate this idea.

Although there is uncertainty regarding the inflammatory drivers of disease among non-T2 forms of asthma, more abundant IFN-γ gene expression, Th1 cells, and secreted mediators of Th1 inflammation have been reported in airway tissues from subsets of asthma patients (*13–17*), suggesting a possible T1-high asthma endotype. The presence of IFN-γ-driven T1 airway inflammation is associated with neutrophilic airway inflammation, itself a marker of corticosteroid-resistant, severe asthma (*6, 13*). Supporting a genetic predisposition towards T1 inflammation, several studies have identified variants in T1 pathway genes (*IL12A*, *IL12RB1*, *STAT4*, and *IRF2*) associated with airflow obstruction and asthma severity (*18*). Respiratory viral infections, the primary trigger of acute exacerbations, induce airway IFN-γ and Th1 responses, raising the possibility that persistent viral illnesses or sub-clinical viral infections may underlie expression of T1 airway inflammation in subsets of asthma patients. Supporting this possibility is the finding that 80% of children with asthma who have high expression of interferon-stimulated genes in their nasal airway epithelium also asymptomatically carry common respiratory viruses in their nasal airway (*19*). Additional questions are raised by recent reports that both T1 and T2 airway inflammation are upregulated in some patients, (*20*) despite the traditional viewpoint that T1 and T2 inflammatory pathways are counter-regulatory (*21*). This possibility is supported by studies showing concomitant airway eosinophilia (T2 marker) and neutrophilia (T1 marker) in subgroups of patients characterized by more severe asthma (*2, 5, 16, 22*).

The Severe Asthma Research Program (SARP)-3 is a US-based, multicenter, longitudinal cohort study of molecular phenotypes of asthma in which patients with asthma, enriched for those with severe disease, undergo deep phenotyping at baseline and longitudinally for at least 3 years. The SARP-3 protocol includes repeated collections of induced sputum cells (processed for cell counts and RNA isolation) and a corticosteroid response test (systemic administration of triamcinolone acetonide). Here, using the SARP-3 sputum cell RNA resource, we generated whole-transcriptome gene expression to determine the frequency, clinical features (including corticosteroid responsiveness), and longitudinal behavior of T1-high, T2-high, and T1/T2-high asthma endotypes. We also leveraged viral metagenomic analysis of these RNA-seq data to determine the frequency of subclinical viral airway infection and the relationships between infection and T1 and T2 immune responses.

## RESULTS

### The frequency, intersection, and clinical characteristics of patients with airway T1-high and T2-high asthma endotypes

Whole transcriptome sequencing data were generated on 782 sputum cell samples collected from 37 healthy adults and 347 adults with asthma in the SARP study cohort (table S1). Of the 745 asthma samples, 260 were collected from patients at their pre-steroid baseline study evaluation visit, where a triamcinolone acetonide (TA) injection was administered, 174 were collected 1-3 weeks later, at the post-TA evaluation visit, and the remainder were collected at annual follow-up visits (Fig. 1). Using transcriptome data from all 782 samples in a weighted gene co-expression network analysis (WGCNA), we identified 42 co-expressed gene networks, which were functionally annotated through pathway, gene ontology, and cell-type enrichment analyses (fig S2). Among these were T1 (65 genes) and T2 (48 genes) inflammation networks (Fig. 2A, 2B). The T1 network was strongly enriched for marker genes of CD8 T cells, natural killer (NK) cells, and T helper 1 (Th1) cells and contained the Th1 master transcription factor gene (*TBX21*), interferon gamma gene (*IFNG*), chemotactic factor RANTES (*CCL5*), and genes critical to interferon signaling (*STAT1*, *CXCL9*, *CXCL10*, *CXCL11*, Fig. 2A). The T2 network was enriched for marker genes of eosinophils, basophils/mast cells, and T Helper 2 (Th2) cells. Moreover, the T2 network included *IL4*, *IL5*, *IL13*, the IL-33 signaling receptor gene (*IL1RL1*), eosinophil transcription factor genes (*GATA1*, *GATA2*, *CEBPE*), and the eosinophil chemotactic gene, *CCL26* (Fig. 2B).

**Fig. 1.**
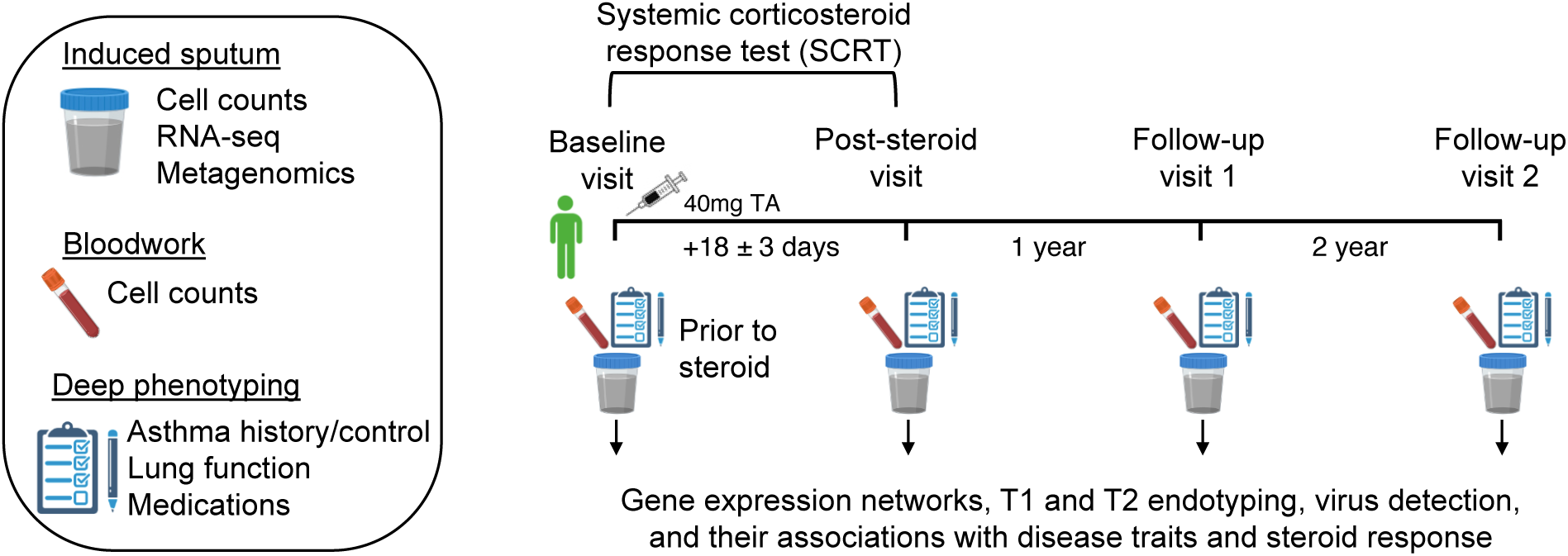
Sampling design for this study. Adults with and without severe asthma yield blood, sputum, and deep phenotyping samples at baseline and follow-up visits, such that inflammatory endotype expression and its associations with disease traits and steroid response can be investigated.

**Fig. 2.**
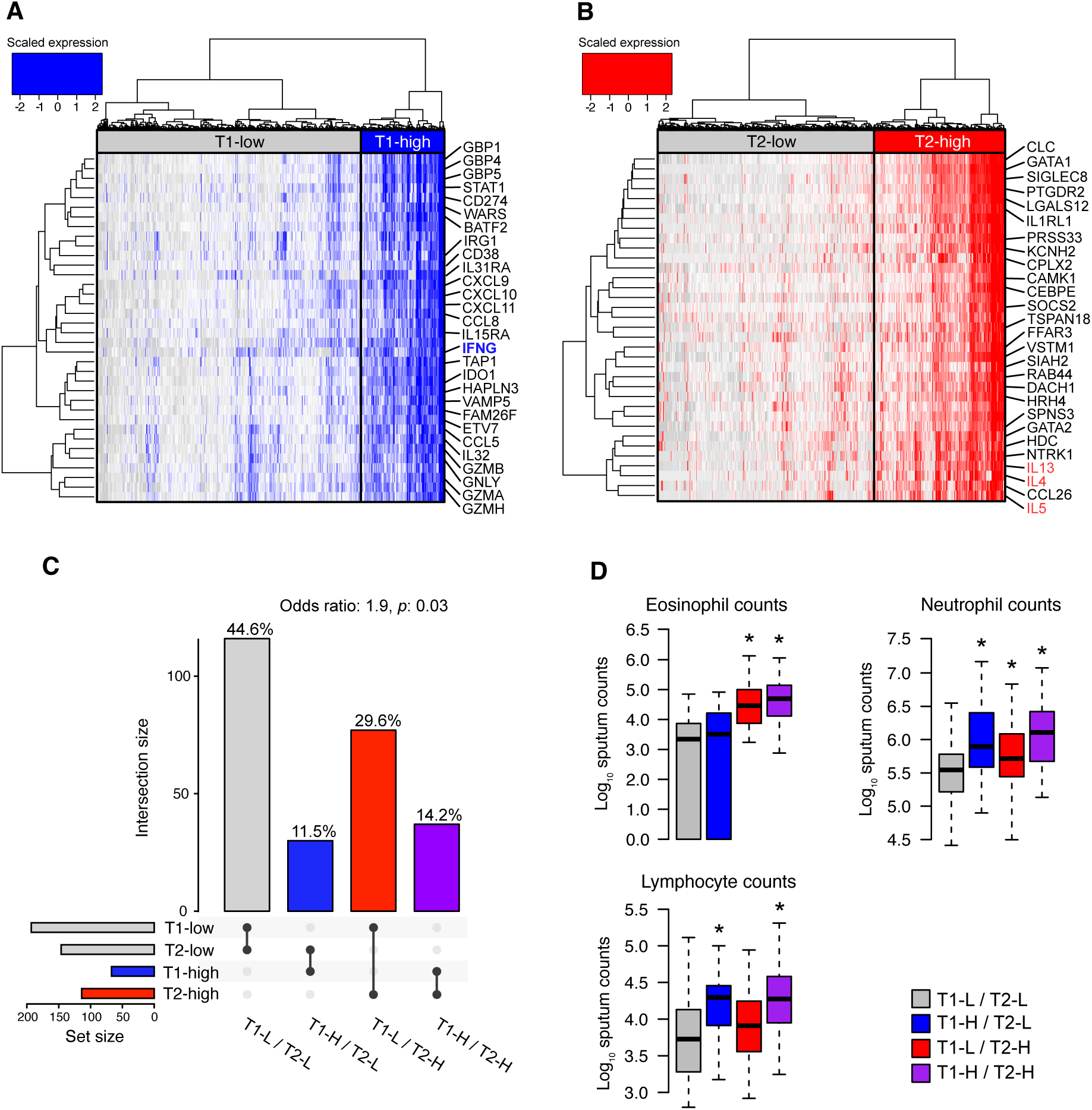
The frequency, intersection, and clinical characteristics of patients with airway T1-high and T2-high asthma endotypes. (**A**) A heat map of 36 T1 network genes as they are expressed among 782 SARP samples hierarchically clustered into T1-low (T1-L) and T1-high (T1-H) groups. A key cytokine of T1 inflammation, IFNG, is highlighted. (**B**) Similar to panel A, but based on 35 T2 network genes. Key cytokines of T2 inflammation, IL-4, IL-5, and IL-13, are highlighted. (**C**) An Upset plot visualizing the intersection between baseline T1 and T2 status among 260 participants, where the number samples in each of the four dual endotype groups is on the y-axis and percent of samples in each group is given over each bar. T2-high endotype status conferred 1.9-fold increased risk of also being T1-high. (**D**) Box plots depicting elevated immune cell counts (log_10_) in the sputum of individuals sampled at baseline (N = 259) who also exhibit T1 and/or T2 inflammatory endotypes (from left to right within a plot, N=115, 30, 77, and 37). Asterisks indicate when log_10_ immune cell counts significantly differ from the dual T1 and T2-low group (p < 0.05) based on an ANOVA that accounts for gender, age, and ethnicity (*p*-values, from left to right: 1.09×10^-10^, 5.98×10^-07^, 2.78×10^-05^, 5.83×10^-04^, 2.13×10^-07^, 0.0122, 0.0250). Data beyond the end of whiskers are not shown.

To determine the frequency of patient subgroups with high expression of T1 and T2 networks at baseline, we independently clustered all sputum samples using expression of the T1 and T2 network genes. We found that 26% of patients had high expression of T1 network genes (T1-high) and 44% were T2-high (Fig. 2A, 2B). Examining the intersection of these classifications, 14.2% of patients were dual-inflamed (T1-H/T2- H), and T2-high endotype status was associated with 1.9-fold increased risk of also being T1-high (p=0.03, Fig. 2C). In addition, we found that 29.6% of patients were T2-high alone (T1-L/T2-H), 11.5% were T1- high alone (T1-H/T2-L), and 44.6% of patients were neither T1-high or T2-high (T1-L/T2-L) (Fig. 2C). Blood and sputum eosinophils were higher in T2-high than in T2-low patients (Fig. 2D, fig. S3). In contrast, sputum neutrophil and lymphocyte numbers (but not blood numbers of these cells) were higher in T1-high patients than in T1-low patients (Fig. 2D, fig. S3). Dual-inflamed patients exhibited sputum cell profiles reflective of both T1 and T2 inflammation, with elevated numbers of eosinophils, neutrophils, and lymphocytes, relative to T1-L/T1-L patients (Fig. 2D).

Both T1-high and T2-high groups were older than the non-inflamed group (table 1). T2-H patients tended to have greater disease severity, as reflected in measures of lung function, medication use (including oral and inhaled corticosteroid), and exacerbation history, than the T1-H patients. However, the T1-H/T2-H patients had the worst asthma severity by these criteria (table 1). These T1-H/T2-H patients were also the oldest and had the latest age of asthma onset.

**Table 1.**
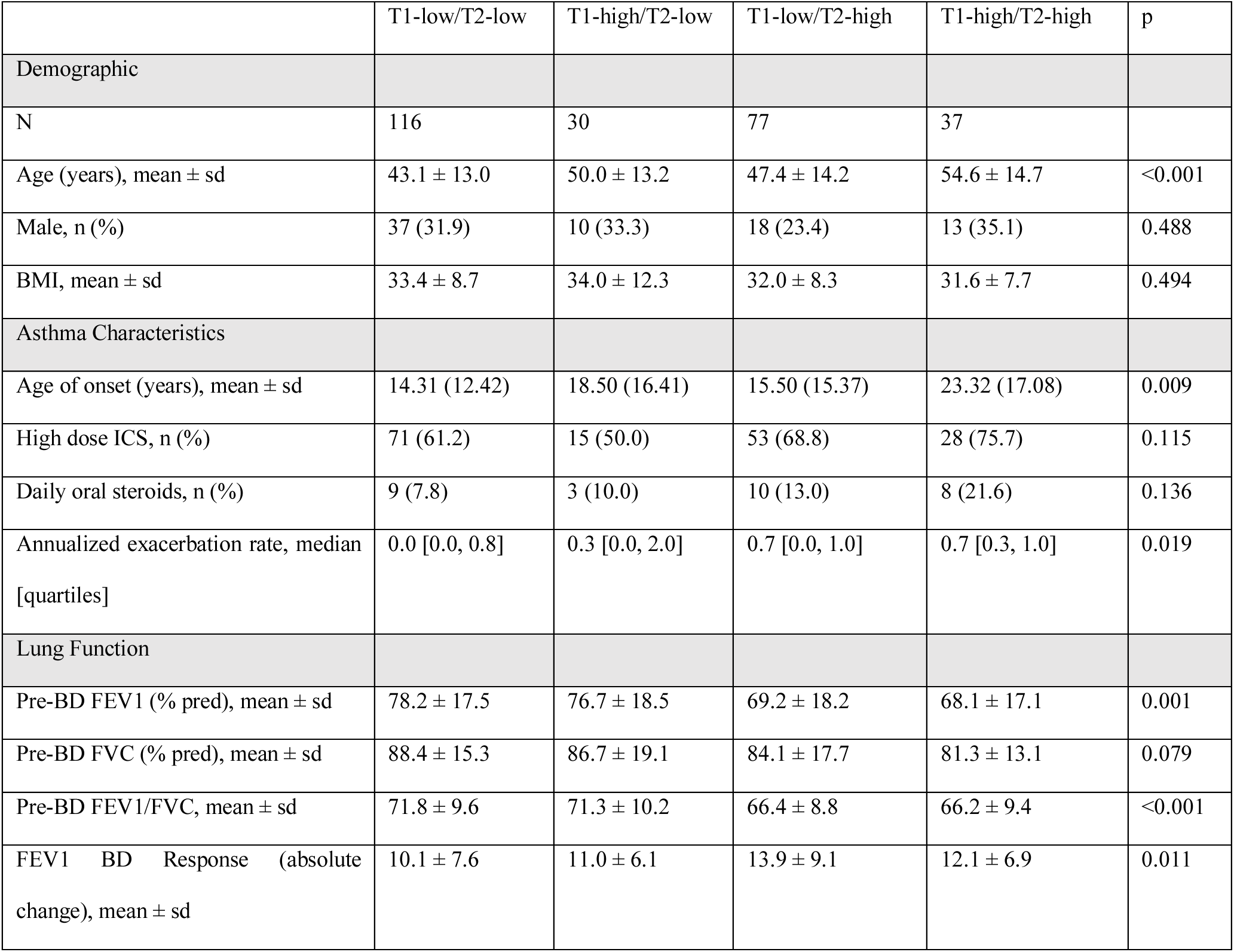
Summary of variation in demographic, asthma, and lung function traits across baseline T1/T2 endotype groups within Severe Asthma Research Program (SARP) participants.

|  | T1-low/T2-low | T1-high/T2-low | T1-low/T2-high | T1-high/T2-high | p |
| --- | --- | --- | --- | --- | --- |
| **Demographic** |  |  |  |  |  |
| N | 116 | 30 | 77 | 37 |  |
| Age (years), mean ± sd | 43.1 ± 13.0 | 50.0 ± 13.2 | 47.4 ± 14.2 | 54.6 ± 14.7 | <0.001 |
| Male, n (%) | 37 (31.9) | 10 (33.3) | 18 (23.4) | 13 (35.1) | 0.488 |
| BMI, mean ± sd | 33.4 ± 8.7 | 34.0 ± 12.3 | 32.0 ± 8.3 | 31.6 ± 7.7 | 0.494 |
| **Asthma Characteristics** |  |  |  |  |  |
| Age of onset (years), mean ± sd | 14.31 (12.42) | 18.50 (16.41) | 15.50 (15.37) | 23.32 (17.08) | 0.009 |
| High dose ICS, n (%) | 71 (61.2) | 15 (50.0) | 53 (68.8) | 28 (75.7) | 0.115 |
| Daily oral steroids, n (%) | 9 (7.8) | 3 (10.0) | 10 (13.0) | 8 (21.6) | 0.136 |
| Annualized exacerbation rate, median [quartiles] | 0.0 [0.0, 0.8] | 0.3 [0.0, 2.0] | 0.7 [0.0, 1.0] | 0.7 [0.3, 1.0] | 0.019 |
| **Lung Function** |  |  |  |  |  |
| Pre-BD FEV1 (% pred), mean ± sd | 78.2 ± 17.5 | 76.7 ± 18.5 | 69.2 ± 18.2 | 68.1 ± 17.1 | 0.001 |
| Pre-BD FVC (% pred), mean ± sd | 88.4 ± 15.3 | 86.7 ± 19.1 | 84.1 ± 17.7 | 81.3 ± 13.1 | 0.079 |
| Pre-BD FEV1/FVC, mean ± sd | 71.8 ± 9.6 | 71.3 ± 10.2 | 66.4 ± 8.8 | 66.2 ± 9.4 | <0.001 |
| FEV1 BD Response (absolute change), mean ± sd | 10.1 ± 7.6 | 11.0 ± 6.1 | 13.9 ± 9.1 | 12.1 ± 6.9 | 0.011 |

### Coincident T1 inflammation in T2-high patients blunts the FEV1 response to triamcinolone acetonide

To determine if airway T1 or T2 inflammation is suppressed by corticosteroid treatment, we analyzed gene network expression data from sputum cells collected before and 1-3 weeks after intramuscular TA injection in 158 patients (Fig. 1). Among T1-high patients, we found that T1 network expression was not significantly downregulated by TA treatment (Fig. 3A). In contrast, among T2-high patients, T2 network expression was strongly downregulated (Fig. 3B). Examining the dual-endotype groups, we found that the TA-induced decrease in T2 network expression occurred in both T1-L/T2-H and T1-H/T2-H patients (Fig. 3C), but not in T1-L/T2-L or T1-H/T2-L patients (Fig. 3C). In addition, we found a strong linear relationship between T2 network expression pre-treatment and the amount of suppression in expression of the T2 network by TA treatment. Specifically, high T2 network expression levels pre-treatment were associated with larger TA-related decreases in T2 network expression, and this was true for both the T1-L/T2-H and T1-H/T2-H subgroups (Fig. 3D).

**Fig. 3.**
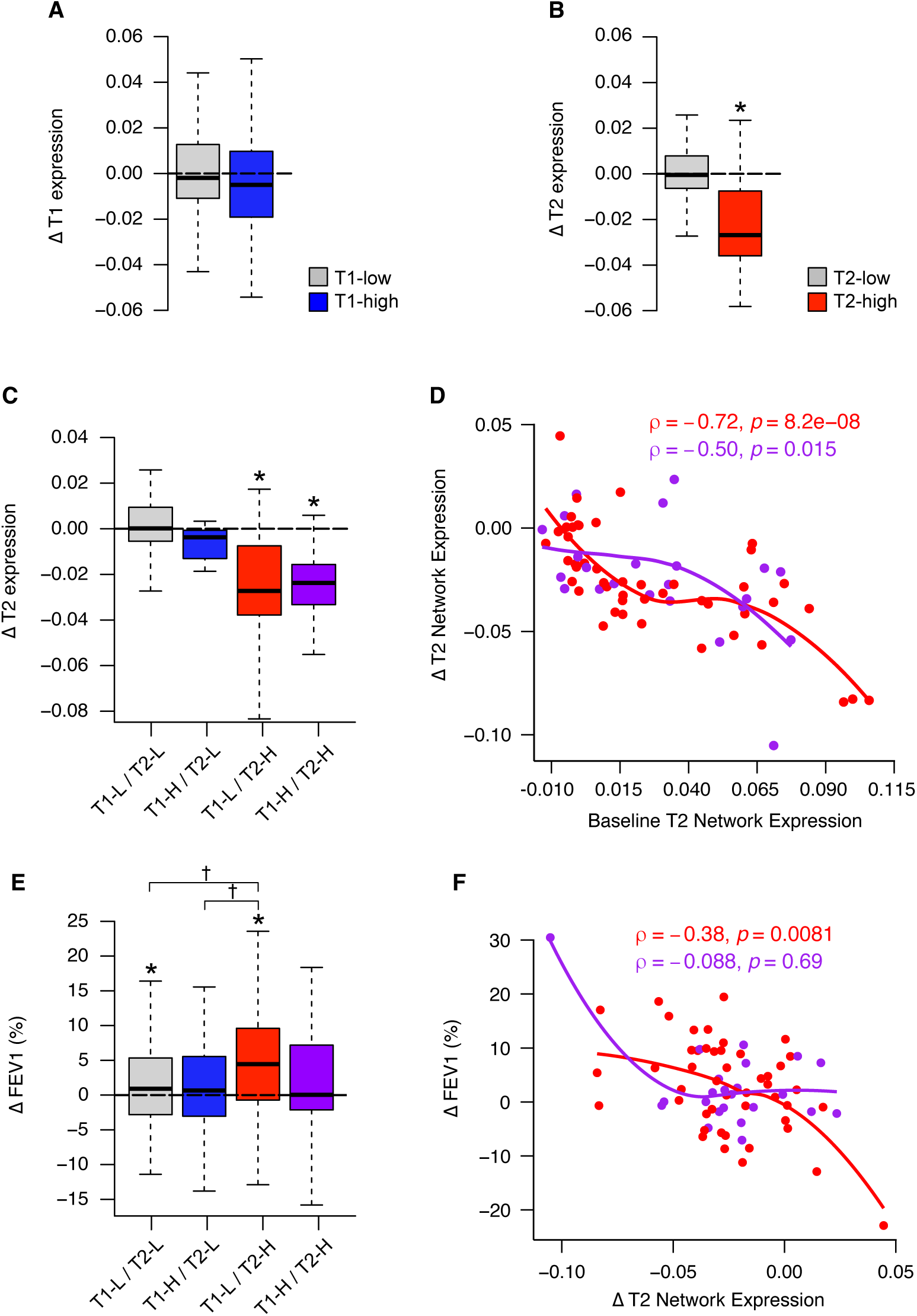
Coincident T1 inflammation in T2-high patients blunts the FEV1 response to triamcinolone acetonide. (**A**) Box plots show patient changes (Δ) in T1 network gene expression from the baseline visit, where steroid was administered, to the post-steroid treatment timepoint, among T1-high and T1-low groups (N=119 for T1-low; N=39 for T1-high). (**B**) Box plots show patient changes (Δ) in T2 network gene expression from baseline to the post-steroid treatment timepoint, among T2-high and T2-low groups (N=88 for T2-low; N=70 for T2-high). The asterisk indicates a significant (*p*-value=1.16×10^-16^) reduction in T2 network expression based on a mixed model that contrasts T2 expression before and after steroid treatment in T2- high individuals, with participant as a random effect. (**C**) Box plots depicting changes in T2 network expression in response to steroid (Δ) among the four, dual T1/T2 endotype groups (from left to right, N=72, 16, 47, and 23). Asterisks indicate significant reduction in T2 network expression based on a mixed model that contrasts T2 expression before and after steroid treatment within a given endotype group (*p*-values, from left to right: 2.35×10^-14^, 1.69e^-07^). (**D**) Plot of the relationship between Δ T2 network expression with steroid against baseline (pre-steroid) T2 network expression for T2-high individuals, stratified by their T1 status, where red = T1-L/T2-H (N=47) and purple = T1-H/T2-H (N=23). Loess curves are overlain on the data points. Spearman correlation coefficients and *p-*values for the two endotype groups are given. (**E**) Box plots depicting patient % FEV1 responses to steroid (Δ) among the dual T1/T2 endotype groups (from left to right, N=116, 30, 77, and 37). Asterisks directly above box plots indicate significant elevation in % FEV1 based on a mixed model that contrasts % FEV1 before and after steroid treatment within a given endotype group (asterisk *p*-values, left to right: 0.0448, 4.42×10^-07^). † symbols between box plots indicate when FEV1 response to steroid is significantly different between groups based on the same mixed model († *p*-values, left to right: 6.48×10^-03^, 0.0324). (**F**) Plot of the relationship between Δ FEV1 with steroid against Δ T2 network expression with steroid for T2-high individuals, stratified by their T1 status, where red = T1-L/T2- H (N=47) and purple = T1-H/T2-H (N=23). Loess curves are overlain on the data points. Spearman correlation coefficients and *p-*values for the two endotype groups are given. For all box plots, data beyond the end of whiskers are not shown.

We next explored whether the size of TA-associated changes in T2 network expression were associated with the magnitude of lung function responses. As noted above, TA decreased T2 network expression in both the T1-L-/T2-H and T1-H/T2-H subgroups, but we found that TA treatment only increased FEV1 in the T1-L/T2-H group, but not in the T1-H/T2-H group) (Fig. 3E). The T1-L/T2-L and T1-H/T2-L subgroups had small or absent FEV1 responses to TA treatment. In addition, TA-induced decreases in T2 inflammation correlated inversely with changes in FEV1 in the T1-L/T2-H group but not in the T1-H/T2- H group (Fig. 3F). Together, these results show that corticosteroids have strong suppressive effects on airway T2 network expression but not on T1 network expression, and that the usual beneficial effects of steroids on lung function in T2-high patients are blunted when they have concomitant T1 inflammation.

### Airway endotype is a patient characteristic with variable expression over time

The ability of TA treatment to suppress airway T2 inflammation suggests that expression of the T2 endotype could vary within patients across time, reflecting changes in treatment, and potentially by other disease-modifying factors or exposures. To investigate this, we examined whether patient T2 endotype at the pre-steroid baseline visit was a significant predictor of T2 status at the 1- (n=113) and/or 3-year follow-up visits (n=111). In logistic regression analyses modeling patient endotype at follow-up visits, we found that patients classified as T2-high at baseline were more likely to be T2-high at follow-up visits (OR = 6.93, p=1.0×10^-8^, fig. S4A). For a subset of these patients (n=72), sputum cell gene expression data were available at all three timepoints, allowing us to examine longitudinal endotype persistence over a three-year period (Fig. 4A). Among 58.3% of patients who were T2-high at least once across the three visits, 31% were T2- high at all timepoints (persistently T2-high), whereas the remaining 69%, variably expressed T2 inflammation. The remaining 41.7% of patients were T2-low at all timepoints. Together, these results strongly suggest that the T2 endotype is a patient characteristic, but that additional factors influence whether a T2-capable patient expresses this endotype at any point in time. We similarly examined evidence for the airway T1 endotype being a patient characteristic. We found that T1-high patients at baseline were more likely to be T1-high at the 1- and 3-year follow-up visits (OR=4.95, p=8.2×10^-6^, fig. S4B). Among those with sputum cell gene expression data at all three timepoints, we found that 52.8% were persistently T1- low (Fig. 4B). Among those who expressed the T1 endotype at least once across this time period, only 11.8% did so persistently, whereas 88.2% were variably T1-high (Fig. 4B). These results indicate that while airway T1 inflammation is also a patient characteristic, endotype expression among predisposed patients is more variable than for airway T2 inflammation.

**Fig. 4.**
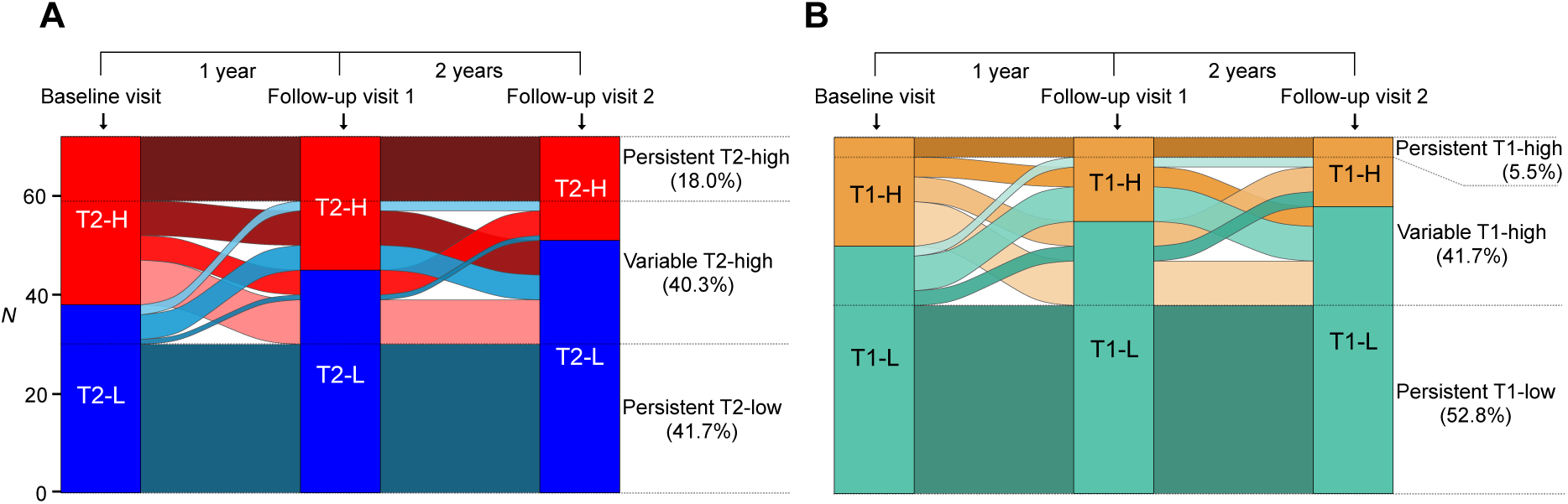
Airway endotype is a patient characteristic with variable expression over time. (**A**) An alluvial diagram depicting patient persistence/variability of T2 status across three time points (N = 72). (**B**) An alluvial diagram depicting patient persistence/variability of T1 status in the same individuals as in panel A across three time points (N = 72).

### Lower airway respiratory virus carriage is associated with expression of the T1 endotype

We next hypothesized that the high within-patient variability in T1 endotype expression is due to periodic respiratory virus carriage. To investigate this, we performed viral metagenomics analysis of the sputum RNA-seq dataset, including only baseline pre-steroid, year 1 and 3 visit, and healthy control samples (605 total samples from 377 participants). We found that 24.3% and 21.6% of asthma and healthy sputum samples, respectively, were positive for respiratory virus reads (Fig. 5A). These viral detections were dominated by human rhinoviruses (A, B, and C species; 47% of detections) and coronaviruses (40% of detections; Fig. 5A, fig. S5A-B). Examination of viral read counts associated with these detections revealed a bi-modal pattern, with virus low and high groups (Fig. 5B). Although T1 network expression was not elevated among the virus-low compared to the non-viral group, it was increased 1.8-fold among the virus-high group (p=1.35×10^-34^, Fig. 5C). Examining the association between T1 endotype status and viral detection groups, we found 20% of T1-high samples were virus-high, versus only 2% of the T1-low samples, an 11-fold increase in the odds of virus-high samples also being T1-high (Fig. 5D). In contrast, T2 network expression was not significantly associated with virus carriage, nor were virus positive samples more likely to be T2-high (fig. S5C-D). Supporting the clinical significance of asymptomatic viral carriage, we found a lower FEV1 in virus-high/T1-high patients than in T1-high patients with low or absent viral carriage (Fig. 5E). Together, these results reveal that lower airway carriage of respiratory viruses is common in adults with asthma and may drive airway T1 inflammation and lower lung function.

**Fig. 5.**
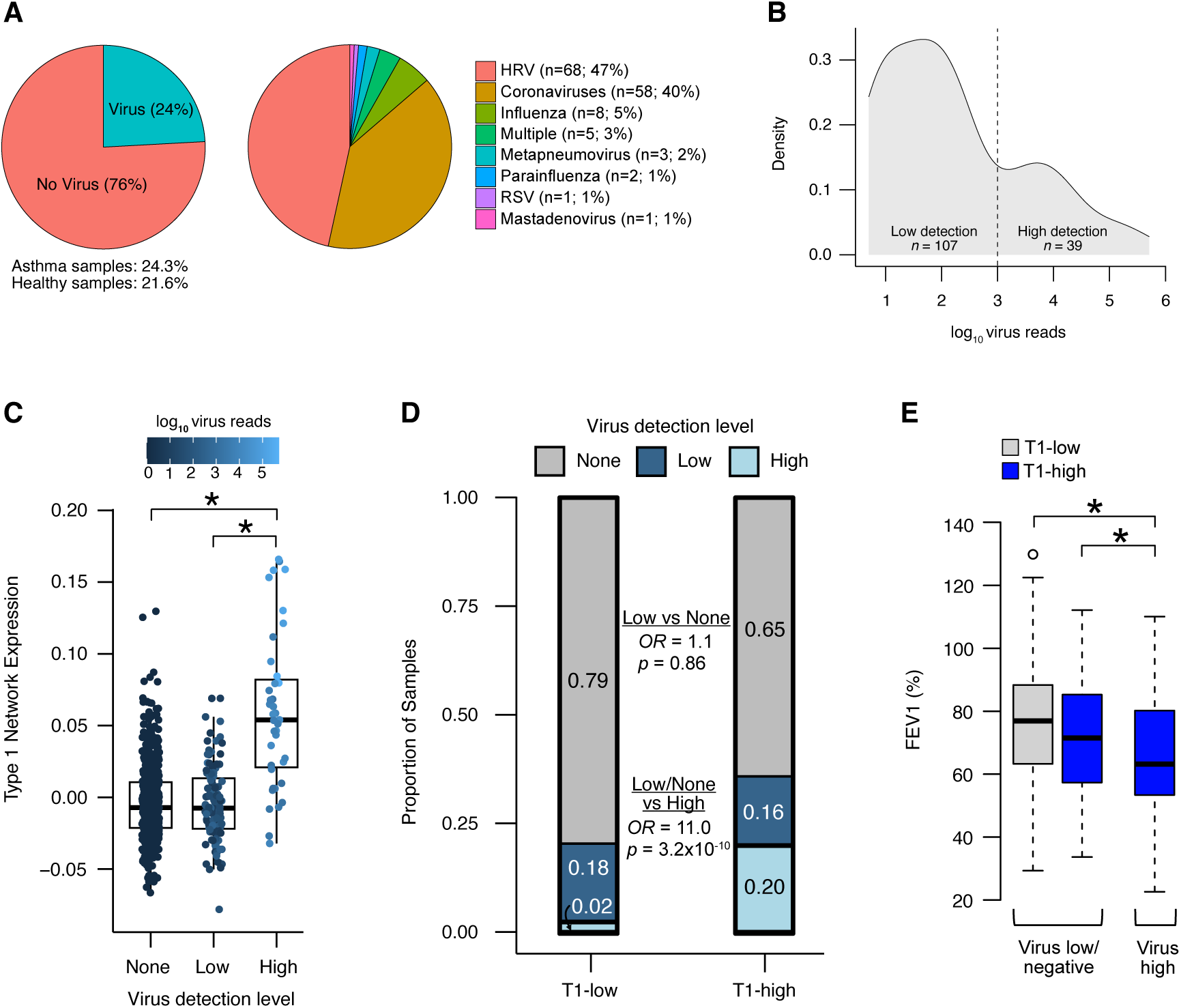
Lower airway respiratory virus carriage is associated with expression of the T1 endotype. (**A**) Pie charts describing the proportion of samples carrying virus as identified using our metagenomics pipeline (left; N=605) or the proportion of different virus types observed among samples carrying virus (right; N=146). (**B**) Density distribution for the log_10_ number of virus reads detected among virus-positive samples (N=146). The dashed line at 1,000 reads marks our threshold to discriminate between “low” and “high” virus detection. (**C**) Box plots visualizing increased T1 network expression among high virus samples (N=39) compared to samples with no (N=459) or low (N=107) virus. Overlaid points are colored by number of log_10_ virus reads in a sample. Asterisks indicate when group differences are significant based on a mixed model predicting T1 network expression as a function of virus status, with participant as a random effect (asterisk *p*-values, left to right: 1.35×10^-34^, 1.30×10^-31^). (**D**) Bar plot depicting the proportion of samples that carry no, low, or high virus within T1-low (N=458) and T1-high (N=147) samples (proportions are listed on each bar). Odds ratios (*OR*) and p-values, describing the excess chance of carrying low virus (compared to no virus; top) or of carrying high virus (compared to no/low virus; bottom) when T1-high compared to T1-low, were calculated using GEE logistic regression. (**E**) Box plots showing lower % FEV1 among T1-high individuals with high virus loads (N=29) compared to individuals who are T1-low (N=423) or who are T1-high while carrying low/no virus (N=116). Asterisks indicate significant differences in FEV1 between groups based on a mixed model with a random effect for participant (*p*-values, from left to right, are 0.0087 and 0.0301).

## DISCUSSION

Here, we leveraged transcriptomic and metagenomic analysis of sputum cells collected during periods of clinical stability over a 3-year period from participants in the Severe Asthma Research Program (SARP)-3 to determine the frequency, persistence, corticosteroid responsiveness, respiratory virus dependence, and associated clinical features of patient subsets with airway inflammation driven by expression of type 1 (T1) and type 2 (T2) cytokines. In particular, we used co-expression network analysis and hierarchical clustering of T1 and T2 gene networks to determine that nearly half of the SARP-3 cohort had T2-high asthma and a quarter had T1-high asthma. Moreover, although traditionally viewed as orthogonal inflammation pathways, we found that 14% of patients exhibited both T1 and T2 airway inflammation. Patients with T2- high asthma tended to have more severe asthma than patients with T1-high asthma, but patients with a dual T1-H/T2-H endotype had the most severe disease. Thus, upregulation of T1 and T2 network expression in the airway has additive effects on asthma severity and fully controlling asthma in these patients may require drugs that inhibit both T1 and T2 cytokine pathways. Notably, T2-high patients at baseline were nearly 7- fold more likely to be T2-high at subsequent timepoints, revealing a patient-level propensity for airway T2 inflammation. This propensity is likely “wired” into a patient’s airway through the presence of a T2 inflammatory cell milieu (e.g. Th2 lymphocytes, eosinophils, mast cells) with enhanced expression of T2 pathway genes, which may be genetically determined (*11, 12*). However, only 38% of patients with T2- high asthma at baseline were persistently T2-high over time. Therefore, for patients who variably express the T2-high endotype, it may be that genetic factors establish the cellular/molecular T2 “wiring,” but additional environmental factors (e.g. aeroallergen exposures) are necessary to periodically activate this pathway. Likewise, we hypothesize that those with persistently T2-high disease experience constant exposure to environmental aeroallergens that maintains their airway T2 inflammation. Alternatively, the airway of these patients may have acquired “inflammatory memory,” through epigenetic programming, leading to a persistent basal inflammation tone or a reduced threshold for activation of airway cytokine production, as has been observed in other disease/tissue contexts (*23, 24*).

We provide strong evidence for the existence of a clinically consequential T1-high asthma endotype. Here, rather than define T1 inflammation based on a single gene or predefined gene set, we used co-expression network analysis to agnostically uncover the gene structure of airway T1 inflammation. These T1 network genes strongly suggest that IFN-*γ*-producing Th1 and CD8+ T cells are central to airway T1 inflammation, as other studies have suggested (*17*). The T1 network also contains IFN-inducible Th1 chemokines (*CCL5* and *CXCL9*-*11*) that recruit Th1 cells to the airway (*25–27*). The patients classified as T1-high were older, prone to exacerbation, and had neutrophilic and lymphocytic inflammation in their airways. Importantly, corticosteroid treatment did not suppress T1 network genes or improve lung function in T1-high patients. Also, the usual beneficial effects of steroids on lung function in T2-high patients was blunted by concomitant T1 inflammation (i.e., in T1-H/T2-H patients). Thus, heterogeneity in the efficacy of corticosteroid treatment in T2-high forms of asthma may be explained by concomitant T1 inflammation. Notably, subsets of T2-high asthma patients are non-responsive to therapeutic proteins targeting the type 2 inflammation cascade (*28, 29*), and it may be that concomitant T1 inflammation also blunts the efficacy of these biologic treatments. Together, our data provide rationale for consideration of drugs that block the T1 inflammation cascade as a strategy to improve disease control in patients with T1-high asthma and in those with a dual endotype (T1-high/T2 asthma).

Our data suggest that T1 inflammation is a patient disease characteristic, as those classified as T1-high at baseline were nearly 5-fold more likely to be T1-high at follow-up visits. However, similar to T2-high patients, we find this propensity towards T1 inflammation varies within these patients over time. Metagenomic analysis of the sputum RNA-seq dataset found that the expression of T1 inflammation was related to carriage of viral RNA for common respiratory viruses, including human rhinoviruses and coronaviruses, because patients with high viral RNA levels were 11-fold more likely to have a T1-high asthma endotype. Importantly, the sputum cell samples analyzed here were collected when patients were free from symptomatic upper or lower respiratory tract infections. This result suggests that a subset of asthmatics “silently” carry respiratory viruses in their lower airways, leading to the activation of T1 inflammation. Supporting this, we have previously observed “silent” nasal carriage of common respiratory viruses in children and that carriage was associated with high airway expression of an interferon-stimulated gene signature (*30, 31*). Moreover, the concept of asymptomatic viral carriage has gained ground during the SARS-CoV-2 pandemic, as increased surveillance for respiratory viruses has revealed the high prevalence of asymptomatic nasal carriage of viruses (*32, 33*). However, our data here represent the first demonstration of widespread asymptomatic viral carriage in the lower airways and raise the possibility that viral carriage drives persistence of airway inflammation. Our study cannot determine if the virus detection represents true asymptomatic viral carriage or a failure to clear illness-associated virus. And despite the high risk of T1 inflammation conferred by viral carriage, we also found that 80% of T1-high patients did not carry a virus, indicating that other unknown factors, potentially bacterial dysbiosis, may underlie T1 gene expression.

In summary, our data reveal independent and interactive effects of T1 and T2 cytokine expression on asthma severity and treatment responses. The longitudinal behavior of these inflammatory patterns indicates that T1-high and T2-high endotypes are patient characteristics modulated by environmental exposures, including respiratory viruses. Patients with T1-high asthma have high unmet treatment needs that may be addressed by targeting the T1/IFN-g inflammation cascade or the respiratory viruses that trigger this cascade in a subset of T1-high patients.

## MATERIALS AND METHODS

### Experimental design

To investigate molecular mechanisms of asthma, we leveraged sputum cell biospecimens from deeply phenotyped participants in the Severe Asthma Research Program (SARP)-3 (*34*). These sputum samples had been collected at baseline characterization visits, before and 1-3 weeks (13 ± 6 days) after a triamcinolone acetonide injection, and at post-baseline follow-up visits at years 1 and 3 (Fig. 1). RNA extracted from these sputum cells was used to generate polyA-selected, whole transcriptome RNA-seq data. These RNA-seq data were used to identify co-expression networks and to detect viral transcripts. The SARP-3 database provided detailed information about the sputum donors’ demographics, lung function, and asthma control as well as information related to treatment response to a single dose of intramuscular triamcinolone acetonide (40mg).

### Participants

Adult patients with asthma were recruited to SARP-3 between Nov 1, 2012, and Oct 1, 2014 by seven clinical research centers in the USA, and 60% had severe disease as defined by the European Respiratory Society/American Thoracic Society (ERS/ATS) criteria (*35*). For this study, we used data collected from 347 SARP-3 asthma participants sampled across different time points (Fig. 1). Selection of samples for RNA-seq was performed to allow robust investigation of baseline endotypes, prioritizing patients with either or both of corticosteroid follow-up and regular longitudinal follow-up samples. Specifically, we included samples from 260 participants at the pre-steroid baseline visit, samples from 174 participants collected 1-3 weeks following triamcinolone acetonide injection (post-steroid visit) (158 of these participants had matched baseline sampling), samples collected from 122 participants 1 year after baseline (113 with matched baseline sampling), and samples collected from 186 participants 3 years after baseline (111 with matched baseline sampling). Three participants with additional interim sampling were collected 2 years after baseline (all three with matched baseline sampling). We also assayed samples from 37 healthy controls with no history of pulmonary/atopic disease or allergic rhinitis. Detailed clinical characterization carried out at baseline visits included administration of questionnaires to capture asthma history, medications, and asthma control, as well as measurement of lung function, evaluation of responses to beta adrenergic agonists and corticosteroids, and collection of computed tomography lung scans and samples of venous blood and induced sputum. Select characterization tests were repeated at regular intervals for three years or longer. Prior SARP-3 publications have previously provided detailed descriptions of the network’s methods and procedures (*5, 6, 34, 36*).

### Systemic corticosteroid response test (SCRT) and maximum bronchodilator reversibility testing (MBRT)

The systemic corticosteroid response test (SCRT) in SARP-3 involved an intramuscular injection of triamcinolone acetonide (TA) (40mg) at the baseline characterization visit followed by a visit 1-3 weeks later to assess TA response (*36*). Maximum bronchodilator reversibility testing (MBRT) in SARP-3 involved measures of spirometry after up to 720 ug of inhaled albuterol and was performed at baseline visits and at each annual in-person visit thereafter (*34*).

### Sputum induction and analysis of induced sputum

Induced sputum was processed within one hour and a portion of sputum (homogenized in a 10% solution of Sputolysin [EMD Millipore, Temecula, Calif]) was used to determine total and differential cell counts (*5*). Homogenized sputum was then centrifuged to generate a cell pellet which was suspended in 1 mL of Qiagen RNAprotect Saliva Reagent. RNA extraction from sputum cells was done by the SARP-3 sputum cell RNA core (UCSF center) using RNeasy Qiagen kits (Qiagen) following an initial DNA elimination step with a Qiagen gDNA elimination column.

### RNA-sequencing of sputum samples

RNA normalization, library preparation, and pooling were performed using the Beckman Coulter Biomek FX^P^ automation system (Beckman Coulter, Fullerton, CA). Total RNA from 886 SARP sputum samples was used to construct whole transcriptome libraries using the KAPA Stranded mRNA-seq library kit (Roche Sequencing and Life Science, Kapa Biosystems, Wilmington, MA) from 20ng of total input RNA, according to manufacturer’s protocols. Barcoded libraries were pooled and sequenced using 125bp paired-end reads on the Illumina HiSeq 2500 system (Illumina, San Diego, CA). Sequence processing and QC, described in the fig. S1 and Supplementary Materials and Methods, resulted in sequence data from 782 samples collected from 384 participants.

### Virus detection using metagenomics

Virus infections were identified and quantified using an in-house meta-transcriptomic NGS pipeline (*30*). This pipeline implements a classic metagenomic approach comprising assembly, binning, classification, and quantification, adding interleaved depletion of human genomic content between each step. A sample was considered positive for a virus if the recovered genome scaffold comprised at least 200 fully determined base pairs and was covered by at least 4 reads.

Only samples from healthy controls and asthma samples from baseline and the two main follow-up visits were used for the viral detection analysis (N=605).

### Defining T1 and T2 endotypes based on sputum co-expression networks

All 782 samples were assigned to “high” and “low” T1 and T2 endotype groups based on hierarchical clustering of genes within the T1 gene network (e.g., *STAT1*, *IFNG*, *CD8A*, *CD8B*; 65 genes total) and the T2 gene network (e.g., *IL4*, *IL13*, *IL1RL1*, *CCL26*; 48 genes total), identified using weighted gene co-expression network analysis (WGCNA) (*37*). See Supplementary Materials for full details of this analysis.

### Statistical analysis

To test for differences in variables among endotype groups at baseline in Table 1 we used ANOVA (normally distributed continuous outcomes), Kruskal-Wallis tests (non-normal continuous outcomes), and chi-squared tests (categorical outcomes). To test for differences in immune cell counts among endotype groups at baseline (Fig. 2D, fig. S3), we used ANOVA, accounting for age, gender, and ethnicity, where ethnicity was binned into “white” and “non-white” due to statistical constraints stemming from small sample sizes of non-white sub-categories. When modeling multiple sample points from a single participant, we used mixed models to account for repeated measures using the lmerTest R package. To estimate the odds of endotype persistence (fig. S4) or of carrying virus in endotype-high versus endotype-low samples (Fig. 5D, fig. S5), we used generalized estimating equations (GEE) logistic regression, allowing us to account for repeat measures within a participant. When only baseline samples were used, odds ratios and p-values from contingency tables were obtained from two-sided Fisher’s exact tests.

## Data Availability

Participant metadata used in this study and gene lists and their enriched pathways for 42 WGCNA co-expression networks can be accessed from https://github.com/seiboldlab/SARP_T1_T2_inflammation. All RNA-seq data in the study will be deposited at Gene Expression Omnibus (GEO).

## Acknowledgments

The authors thank all the volunteers who participated in these studies. They also thank the investigators and coordinators in the SARP who contributed to this manuscript by recruiting and characterizing participants, collecting biospecimens, and coordinating data collection and analysis. The following companies provided financial support for study activities at the Coordinating and Clinical Centers beyond the third year of patient follow-up: AstraZeneca, Boehringer-Ingelheim, Genentech, GlaxoSmithKline, Sanofi-Genzyme- Regeneron, and TEVA.

## Author Contributions

Conceptualization: JVF, MCP, MAS; Methodology: JVF, MCP, MAS; Formal analysis: PGW, NDJ, SPS, MCP, JVF, EP, MT, MG, CMM, MAS; Investigation: JLE, CR; Resources & Supervision: JVF, SEW, ERB, MC, SAC, SCE, ATH, WM, EI, BDL, LD, NNJ, MWJ, KS; Data curation: MAS, NDJ, DTM, BRP Writing – original draft: JVF, MAS; Writing – review & editing: JVF, MCP, MAS, PGW, NDJ, SPS, EP, MT, MG, CMM, JLE, CR, SEW, ERB, MC, SAC, SCE, ATH, WM, EI, BDL, LD, NNJ, MWJ, KS, DTM, BRP.

## Data and material availability

Participant metadata used in this study and gene lists and their enriched pathways for 42 WGCNA co-expression networks can be accessed from https://github.com/seiboldlab/SARP_T1_T2_inflammation. All RNA-seq data in the study are deposited at Gene Expression Omnibus (GEO) under accession number TBD.

