## Supplementary Materials for "Asthma severity and corticosteroid response depend on variable type 1 and type 2 inflammation in the airway"

### Supplementary Materials and Methods

*Processing of RNA-seq data*

Raw sequence reads were trimmed using skewer v0.2.2 (end-quality 15, mean-quality 25, min 30) (*1*) and then aligned to the human reference genome (GRCh38) using HISAT2 v2.1.0 (*2*) with default parameter settings. Gene quantification was performed using htseq-count (mode=intersection-nonempty, stranded=reverse, a=20) (*3*) with GRCh38 Ensembl v84. Samples were filtered out based on standard QC parameters (number of mapped reads > 15 million, % reads to mitochondrial genes < 40), resulting in RNA-seq data for 782 samples collected from 384 participants (347 with asthma) across the different time points.

*Defining T1 and T2 endotypes based on sputum co-expression networks*

To cluster genes into co-expressing groups that may correspond to discrete biological function, we applied weighted gene co-expression network analysis (WGCNA) to VST-normalized counts (DESeq2; *4*) for the top 10,000 most variant genes (selected after filtering out ribosomal and mitochondrial genes, unannotated ENSG genes, and genes without 10 or more counts observed in at least 15% of samples). Note that in addition to all 782 samples in the current study, this analysis also included 48 non-SARP asthma samples that are not part of the current study. The analysis was run using soft threshold=16, type=signed, minClusterSize=10, deep split=3, pamStage=FALSE, and pamRespectsDendro=TRUE. This analysis identified 42 networks, which were assigned biological functions based on pathway enrichment analysis using Enrichr (*5*). Among these networks was one strongly enriched in markers of T1 inflammation (e.g., *STAT1*, *IFNG*, *CD8A*, *CD8B*; 65 genes total) and another strongly enriched in markers of T2 inflammation (e.g., *IL4*, *IL13*, *IL1RL1*, *CCL26*; 48 genes total). We categorized the 782 samples into T1- and T2-"high" and "low" groups by hierarchically clustering samples (ward.D2 clustering of a Euclidean distance matrix using *hclust* in R) based on all network genes, for T1 and T2 gene sets in turn, whose expression exhibited a Pearson correlation $\geq 0.7$ with the network overall (i.e., network eigengenes) (36 T1 genes and 35 T2 genes). The first split in the dendrograms were used to distinguish high and low-expressers of each endotype.

### Supplementary Tables

**Supplementary Table 1.** Description of SARP participants in this study.

| | Healthy (n=37) | Asthma SARP (n=347) |
|---|---|---|
| Age (years), mean ± sd | 38.9 ± 13.3 | 47.7 ± 13.8 |
| Male, n (%) | 8 (25.0) | 109 (31.4) |
| BMI, mean ± sd | 25.9 ± 5.6 | 32.4 ± 8.6 |
| Race, n (%) | | |
| White | 17 (53.1) | 230 (66.3) |
| Black | 4 (12.5) | 79 (22.8) |
| Asian | 6 (18.8) | 13 (3.7) |
| Other/Multiple | 4 (12.5) | 17 (6.6) |
| Hispanic, n (%) | 1 (3.1) | 12 (3.5) |
| Pre-BD FEV1 (% pred), mean ± sd | 96.9 ± 9.7 | 73.2 ± 18.2 |
| Pre-BD FVC (% pred), mean ± sd | 98.0 ± 12.2 | 85.3 ± 16.4 |

### Supplementary Figures

**Fig. S1.** Consort-style diagram summarizing samples and subjects sequenced and analyzed in the study.

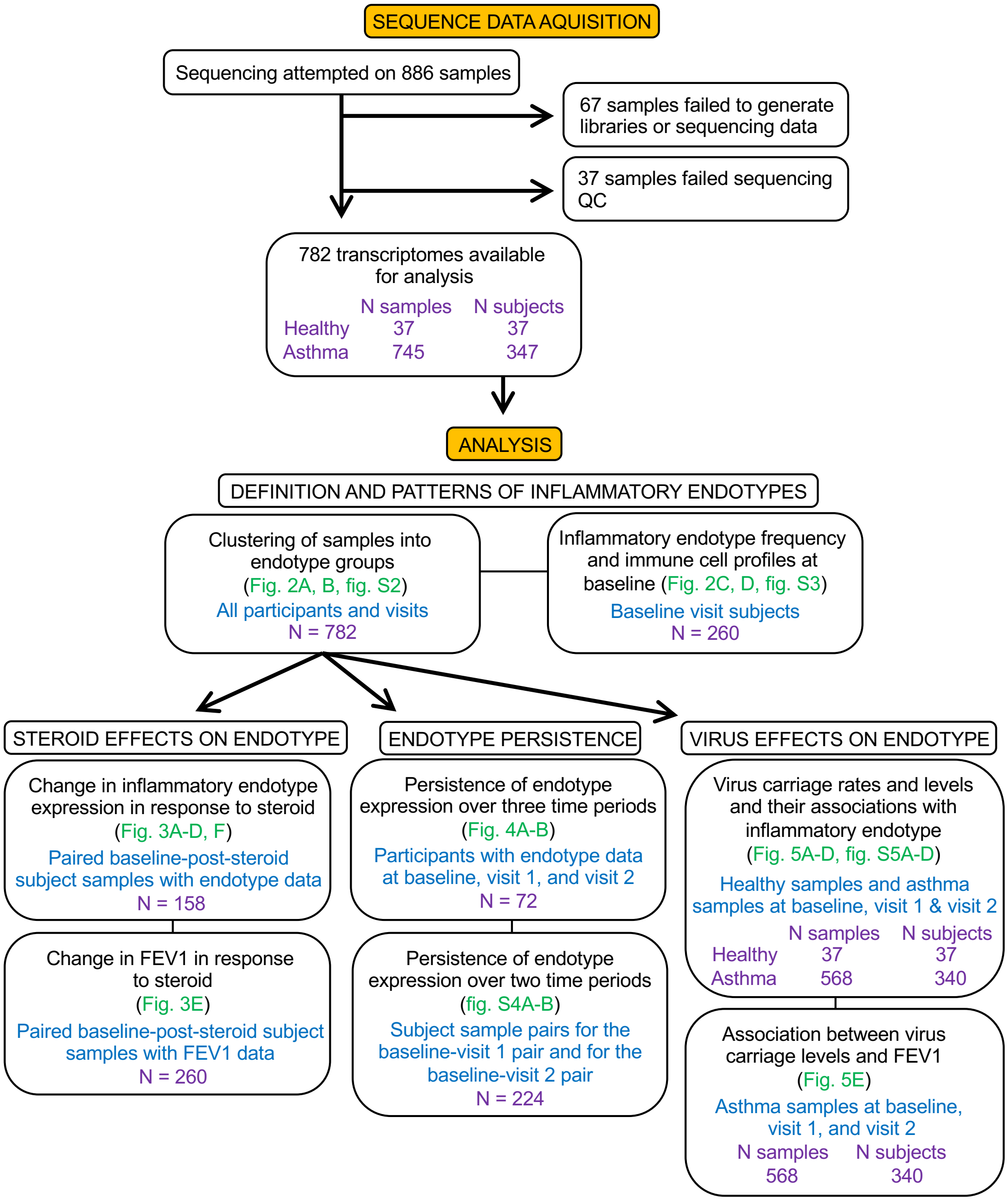

**Fig. S2.** Heat map of pairwise Pearson correlation coefficients among eigenenes for 42 WGCNA gene networks identified from RNA-seq data on 782 SARP samples, enabling visualization of groups of highly correlated networks in the dataset. Identifiable groups, including T1 and T2 inflammation, immune signaling, and epithelial expression, are highlighted. Functional descriptions and key genes for each network are given.

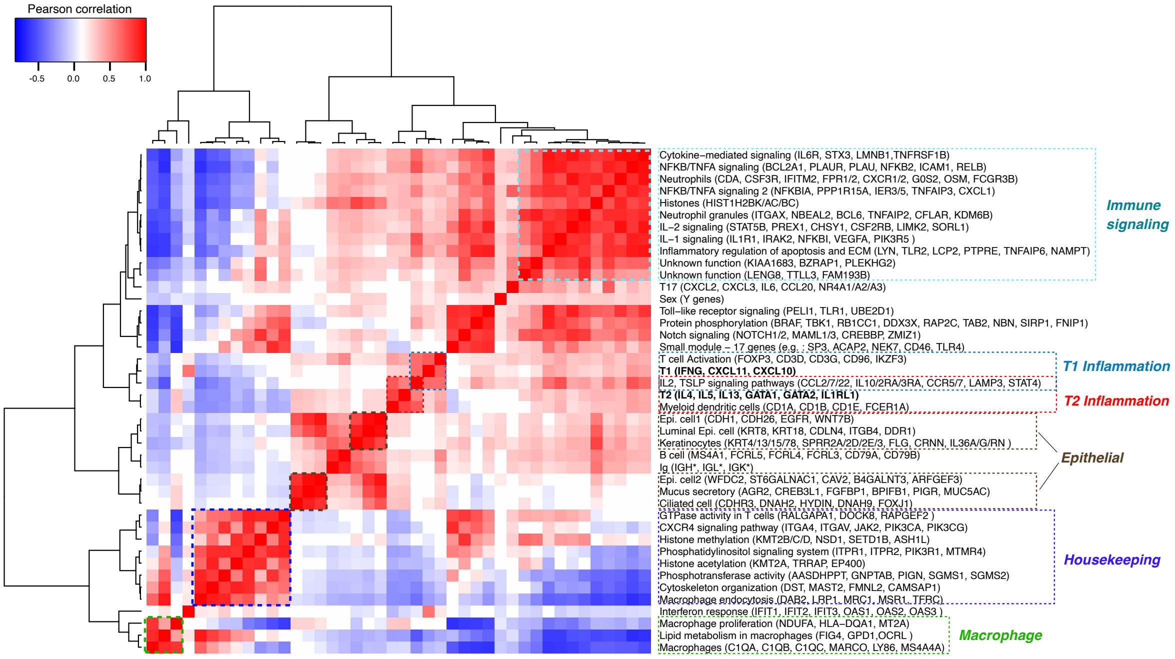

**Fig. S3.** Box plots depicting immune cell counts ($log_{10}$) in the blood of individuals sampled at baseline (N = 260) who also exhibit T1 and/or T2 inflammatory endotypes (from left to right within a plot, N=116, 30, 77, and 37). Asterisks indicate when $log_{10}$ immune cell counts significantly differ from the dual T1 and T2-low group ($p < 0.05$) based on an ANOVA that accounts for gender, age, and ethnicity (*p*-values, from left to right: $4.19 \times 10^{-08}$, $1.73 \times 10^{-04}$). Data beyond the end of whiskers are not shown.

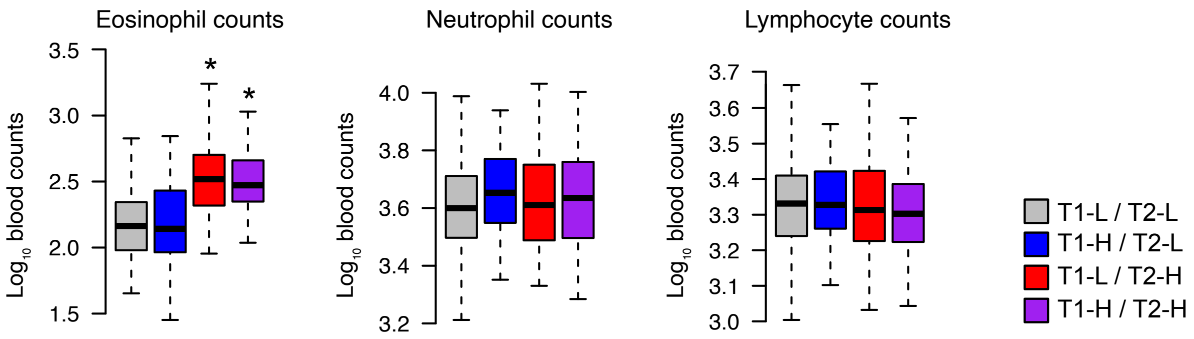

**Fig. S4.** (**A**) An alluvial diagram depicting persistence of T2 status between baseline and 1-year follow-up visits (top; N=113) and between baseline and 3-year follow-up visits (bottom; N=111). The odds ratio (*OR*) and p-value, describing the excess chance that a T2-high individual at follow-up was also T2-high at baseline compared to having switched from T2-low, were estimated using GEE logistic regression, where the two sets of visit pairs were analyzed together. (**B**) The same as panel A, except related to persistence of T1 status.

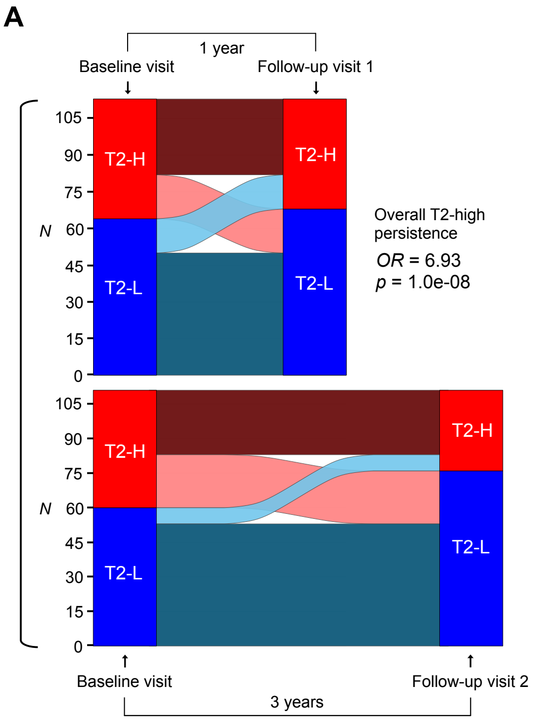

**Fig. S5.** (**A**) A pie chart describing the proportion of each human rhinovirus species identified using our metagenomics pipeline (N=68). (**B**) similar to panel A but for different coronavirus species (N=58). (**C**) Box plots visualizing no difference in T2 network expression among high virus (N=39), low virus (N=107), and no virus (N=459) samples. Effects of virus group were carried out using a mixed model predicting T2 network expression as a function of virus status, with participant as a random effect. Overlaid points are colored by number of $\log_{10}$ virus reads in a sample. (**D**) A bar plot depicting the proportion of samples that carry no, low, or high virus within T2-low (N=359) and T2-high (N=246) samples (proportions are listed on each bar). Odds ratios (*OR*) and p-values, describing the excess chance of carrying low virus (compared to no virus; top) or of carrying high virus (compared to no/low virus; bottom) when T2-high compared to T2-low, were calculated using GEE logistic regression.

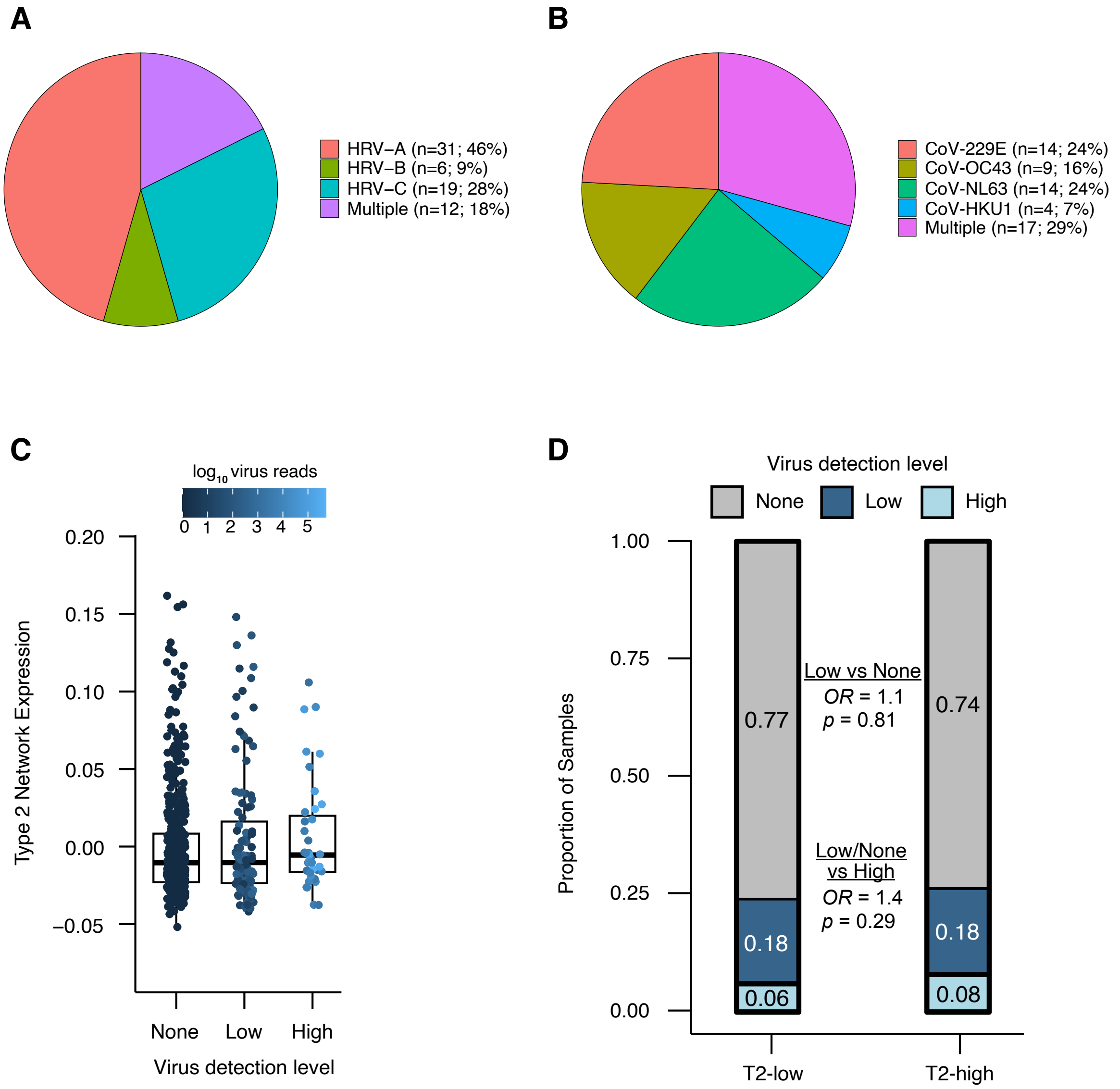
